# Psychotherapies and Psychological Support for Individuals Facing Psychological Distress during the COVID-19 Pandemic: A Scoping Review

**DOI:** 10.1101/2023.09.04.23295013

**Authors:** Mao Yagihashi, Atsushi Sakuma, Michio Murakami

**Author notes:** **Corresponding Author** Mao YAGIHASHI. C209 Techno Alliance, 2-8 Yamadaoka, Suita, Osaka 565-0871, Japan.

## Abstract

In this scoping review, we investigated psychotherapies and psychological support provided the coronavirus disease 2019 (COVID-19) pandemic to clarify its recipients and the methods employed. We used Scopus and PubMed as the search engines on October 18, 2022, employing specific search terms ("COVID*" AND ("Psychotherap*" OR "psychological support*") AND "psychological distress*"). The initial search yielded 153 articles, of which 18 met the eligibility criteria after two rounds of screening. The distribution of participants ranged from the general population to patients with COVID-19 and those who had recovered. However, no studies of patients with post-COVID-19 sequelae were found. The distribution of the types of and psychological support varied and the use of new technology was suggested. Online interventions comprised the majority of the means of psychotherapies and psychological support. This study suggests that psychotherapies and psychological support during the COVID-19 pandemic were influenced by the social situation.

## 1. Introduction

The deterioration of mental health caused by the coronavirus disease 2019 (COVID-19) is a public health concern globally [1]. The exacerbation of psychiatric symptoms amid the COVID-19 spread can be attributed to the bio-psycho-social model [2], as is typically observed in the development of psychiatric symptoms. This model includes biological factors such as physical pain and its sequelae, that is, lingering symptoms caused by COVID-19 infection [3, 4]; behavioral factors such as lockdown and refraining from activities [5]; and psychological factors such as isolation due to behavioral restrictions and fear of infection [6], a sense of self-inflicted guilt for having been infected [7], and discrimination against infected persons or persons with specific backgrounds [8]. Other social factors include economic deprivation caused by lockdowns and business restrictions [9]. The World Health Organization declared the end of the COVID-19 pandemic on May 5, 2023. However, in the future, more attention should be paid to psychological support for those who have recovered from the acute symptoms of COVID-19 but still suffer from chronic symptoms. One of the findings of disaster psychiatry, which focuses on pandemics, is that people may choose to avoid seeking psychological help during the acute phase of an infectious threat, and many mental health disorders will surface over time as the infectious threat decreases [10]. This is because requests for mental healthcare and support services may increase rapidly as the threat of infection declines.

An early review of healthcare workers’ mental health during the COVID-19 pandemic reported six intervention studies on healthcare workers; however, none mentioned their effectiveness [11]. There has also been a review of psychological support interventions for healthcare professionals and informal caregivers (i.e., family members and others close to them) during the COVID-19 pandemic [12]. Additionally, a narrative review of interventions for family caregivers of patients with amyotrophic lateral sclerosis reported their experience using telemedicine [13]. However, as of July 5, 2023, and after conducting another search on Scopus using the terms “COVID,” “distress,” and “psychological support,” we found 21 reviews but no papers that directly addressed our research question. In other words, there are no studies (1) targeting patients or the general population rather than solely healthcare providers or (2) summarizing the types of psychotherapies and psychological support offered to these diverse audiences and the mediums through which they were delivered.

Knowledge of these data would help decide what types of psychotherapies and psychological support to provide to mitigate the psychological distress of those infected not only with COVID-19 but also with other possible infectious diseases in the future. Furthermore, a summary of existing articles on the subject would help us understand global trends and what perspectives are lacking, which would be useful for conducting intervention research in the future. Therefore, we conducted a scoping review to identify the types of psychotherapies and psychological support provided and by whom during the COVID-19 pandemic.

## 2. Methods

This study was conducted according to the Preferred Reporting Items for Systematic Reviews and Meta-Analysis Extension for Scoping Review (PRISMA-ScR) guidelines. The review was conducted in four phases: identification, screening, eligibility assessment, and final synthesis (Figure 1).

**Figure 1.**
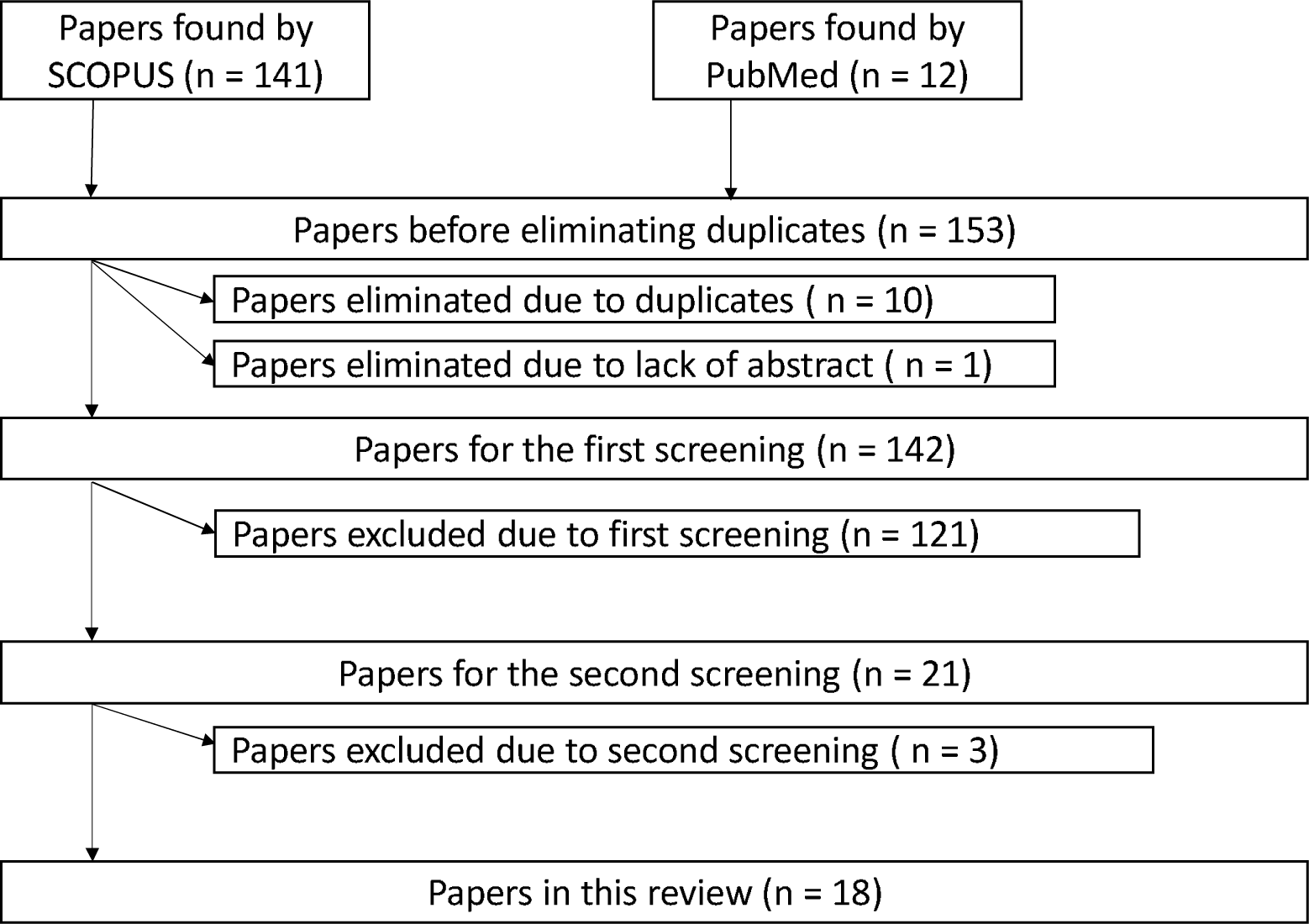
Screening process in this study.

### 2.1 Identification

We conducted our search on October 18, 2022. Since the objective was to determine the types of psychotherapies and psychological support provided for psychological distress during the COVID-19 pandemic, the primary keywords were “COVID-19,” |Psychotherapy” (or) “psychological support,| and “psychological distress.” We searched two article engines (Scopus and PubMed). According to our research question, the principal criterion was whether psychotherapies and psychological support were implemented during the COVID-19 pandemic. Other inclusion criteria were as follows: the article had to be an original paper or review article, its full text had to be available (not just the abstract), and it had to be written in English. Protocol articles were also included among the articles obtained by the search engine but were excluded from the screening. The search formula for Scopus was the following: TITLE-ABS-KEY ("COVID*" AND ("Psychotherap*" OR "Psychological support*") AND "psychological distress*") AND (LIMIT-TO (DOCTYPE,"ar") OR LIMIT-TO (DOCTYPE,"re")) AND (LIMIT-TO (LANGUAGE,"English"))). The search formula for PubMed was as follows: (("covid*"[Title/Abstract] OR "COVID-19"[MeSH Terms]) AND ("psychotherap*"[Title/Abstract] OR "psychological support*"[Title/Abstract]) AND (" Psychological Distress *"[Title/Abstract] OR "Psychological Distress"[MeSH Terms])) AND ((ffrft[Filter]) AND (fha[Filter]) AND (clinicaltrial[Filter] OR meta-analysis[Filter] OR randomizedcontrolledtrial[Filter] OR review[Filter] OR systematicreview[Filter]) AND (fft[Filter])). The search yielded 153 entries (141 from Scopus and 12 from PubMed).

### 2.2 Screening

The screening procedure is illustrated in Figure 1. First, all entries (n = 153) were outputted using Endnote reference management (Endnote X8, Clarivate Analytics). Duplicates (n = 10) and those without abstracts (n = 1) were removed according to the inclusion criteria. In the first screening, two independent experts, M.Y. (major in clinical psychology) and M.M. (major in risk science), determined whether the inclusion or exclusion criteria were met based on the titles and abstracts. The results were integrated through discussion. The kappa coefficient of agreement was 0.68. When the judgments were not in accordance with the two independent experts mentioned above, a third expert, A.S. (major in psychiatry, especially liaison psychiatry), was consulted. A total of 121 papers were excluded in the first screening, while 21 remained eligible.

During the second screening, all 21 papers were manually obtained. Two independent experts (M.M. and A.S.) read half of the articles and M.Y. read them all. We confirmed that they met the inclusion criteria for this study and itemized the content according to the country or region in which psychotherapies and psychological support were provided, the target population for psychotherapies and psychological support, the types and means of psychotherapies and psychological support, and the person who conducted the psychotherapies and psychological support. The details are as follows:

- Types of intervention participants Participants were classified according to the descriptions provided in the articles, allowing for duplicate responses.
- Number of participants The total number of participants and the number of intervention and control groups were recorded according to each study design.
- Countries and regions The countries where the interventions were implemented were documented and categorized based on the descriptions of the articles.
- Types of psychotherapies and psychological support Duplicate responses were allowed and classifications were made broadly considering the real-world settings of interventions. Content in the papers guided the classification process (i.e., psychotherapies and psychological support "based on humanistic psychology" was classified as humanistic psychology). Furthermore, in the case of intervention studies that were divided into control and intervention groups, support for the control group was not included (e.g., distribution of newsletters and provision of regular support).
- Types of intervention trials Duplicate responses were allowed. The following items were classified: validation of feasibility, randomized controlled trials (RCTs), established existing interventions, and others.
- Means of psychotherapies and psychological support Duplicate responses were allowed and classified as remote, face-to-face, and others. Remote communication included both voice-only and video calls but was defined as remote when real-time two-way communication was possible.
- Types of psychotherapists and psychological supporters Duplicate responses were allowed, including doctors, nurses, psychologists/counselors, and others. To maintain consistency, "psychologist" and "counselor" were considered to be identical for classification purposes, acknowledging that the definitions may vary by country.
- Outcomes Duplicate responses were allowed. They were written according to descriptions in the literature.

### 2.3 Eligibility and inclusion

After the second screening, if the two independent researchers disagreed on the results, the three experts discussed and reached a consensus. Consequently, 18 papers were included in this study [14–31]. The paper’s content was compiled by M.Y. and subsequently reviewed and edited by M.M. and A.S.

## 3. Results

Table 1 summarizes the papers included in this review and Figure 2 shows the distribution of each item.

**Table 1.** Overview of the papers included in this study.

| Ref. | Period | Country | Participants | Number of participants; interventions (I) vs. controls (C) | Types of intervention trials | Types of psychotherapies and psychological support | Means of psychotherapies and psychological support | Interventionist/ supporter | Outcome(s) | Paper summary |
| --- | --- | --- | --- | --- | --- | --- | --- | --- | --- | --- |
| [14] | Assessments were between September 2019 and June 2020 | Jordan | Refugees | 410 (I: 204, C: 206) | RCT | Usual care vs. Problem Management Plus | Face-to-face | Trained but non-specialist | PCL<br>HSCL | This study examined the psychological impact of the pandemic on the mental health of Syrian refugees living in the Azraq camp in Jordan. Severe post-traumatic stress disorder symptoms were less among the refugees assessed during the pandemic than those assessed before the pandemic. |
| [3]<br>[15] | Participants were recruited between April 14 and May 30, 2020 | Oman | Public | 46 (I: 22, C: 24) | RCT | Autonomic weekly newsletter containing self-help therapy vs. therapy based on the principles of cognitive behavioral therapy (CBT) and acceptance commitment therapy (ACT) | Remote | Psychologist/counselor | PHQ<br>GAD | This study compared the efficacy of therapist-guided online therapy (CBT + ACT) with self-help therapy by weekly newsletters for people living in Oman during the COVID-19 pandemic. The levels of anxiety and depression reduced in both groups; however, the reduction was higher in the intervention group than in the control group. |
| [16] | Participants were recruited between February 21 and July 31, 2020 | Italy | Patients after recovery from COVID-19 | 264 (I: 16) | Established existing intervention | Supportive psychotherapy based on humanistic psychology | Remote | Psychologist/counselor | HADS<br>IES-R | This study described the psychological status of patients recovering from COVID-19. In particular, it focused on anxiety, depressive symptoms, and post-traumatic stress. The results showed that patients reported anxiety |
|  |  |  |  |  |  |  |  |  |  | (28%), depression (17%), and post-traumatic stress (36.4%). In total, 13.8% of patients received psychological visits and 6.1% were received psychological support. Patients who had recovered from COVID-19 reported negative mental health outcomes in the months following discharge. |
| [17] | Not described in detail and the study just mentioned "the intervention program in times of COVID-19" | Mexico | Public | 34 (I: 34) | Non-experimental, cross-sectional, exploratory-descriptive design | "Psychology for all" support program based on brief psychotherapy and CBT | Remote | Not described | Kessler | This study implemented an intervention program to reduce the aftereffects of the COVID-19 pandemic (implementation of "Psychology for All") and measure mental health outcomes. Pre- and post-implementation comparisons of the implemented intervention program showed a decrease in the value of psychological distress. |
| [18] | Participants were recruited in July and August 2020 | India | Primary caregivers of children with cancer | 100 (I: 22) | Established existing intervention | Supportive psychotherapy based on humanistic psychology | Remote | Psycho-oncologist | PHQ<br>GAD | Psychological distress was prospectively assessed via telephone calls using PHQ-9 and GAD-7 for the primary caregivers of pediatric cancer patients. Among high-risk individuals, GAD scores decreased after the intervention, whereas PHQ scores did not. |
| [19] | Participants were recruited between November 2020 and March 2021 | Italy | Healthcare workers | 225 (I: 68, C: 157) | Established existing intervention | Eye movement desensitization and reprocessing (EMDR) therapy | Remote | Psychologist/counselor | IES-R<br>THERMO | This study investigated post-traumatic stress disorder in a sample of Italian healthcare workers during the COVID-19 outbreak and evaluated the efficacy of EMDR therapy on this population. The results showed that the IES-R and THERMO values decreased after the intervention in a pre- and post-intervention comparison of the EMDR treatment group only. |
| [20] | The details were not described | Canada, UK-based with multiple countries, USA, China | Public, staff, patients with COVID-19 | 32 (Canada), 646 (UK-based with multiple countries), 24 (USA), 235 (China) | Verification of feasibility, RCT, established and existing intervention | Virtual reality | Not described | Not described | Satisfaction etc. | This systematic review evaluated the role of virtual reality as a psychological intervention tool for mental health problems during the COVID-19 pandemic. Virtual reality is a beneficial tool for intervention in individuals with mental health problems. |
| [21] | Participants were enrolled between February 6 and March 9, 2020 | China | Patients with COVID-19 | 67 (I: 67) | Established existing intervention | Simplified CBT-Insomnia (S-CBTI) | Remote (patients were asked to participate in behavioral therapy by mobile phone and paper-based systems) | Medical doctors | ISI | This study developed a brief CBT for insomnia (S-CBTI) in patients with COVID-19 and comorbid insomnia symptoms and tested its efficacy in a self-controlled study. The second objective was to compare the effectiveness of S-CBTI between acute and chronic insomnia in women with COVID-19 and comorbid insomnia symptoms. The ISI scores in the intervention group improved before and after the intervention. After the intervention, the mean ISI score of the acute insomnia group was lower than that of the chronic insomnia group. The decrease in the ISI scores and improvement in sleep duration from baseline to post-intervention was greater in the acute insomnia group than in the chronic insomnia group. |
| [22] | Not described in | Singapore | Healthcare | 80 (I: 40, C: 40) | Established | Mobile app-based | Mobile app | Not described | PCL, DAS, | This study investigated the effects of a |
|  | detail; participants were recruited through a participant database hosted by the medical school |  | workers during the COVID-19 pandemic |  | existing intervention | mindfulness |  |  | FCV-19S, PWI, ProQOL, PSQ, FFMQ, SCS, and digit span Tests (forward and backward) | mindfulness practice delivered using Headspace on the psychological and cognitive outcomes of healthcare workers in Singapore. The intervention improved the fear of COVID-19, compassion satisfaction, trait mindfulness, self-compassion, sleep quality, and forward digit span task at the 1-month follow-up. |
| [23] | Patients with COVID-19 were recruited from June to August 2020 | Indonesia | Patients with COVID-19 | 47 (I: 42) | Verification of feasibility | Video-based psychotherapy including relaxation, management of thoughts and emotions, and mindfulness | Watching videos | The video (the creators were psychiatrists) | SUDS | This study investigated the effectiveness of video-based psychotherapy in reducing distress in COVID-19 patients being treated in an isolation ward. After the intervention, the SUDS scores decreased in the intervention group. |
| [24] | The program ran between March 11, 2019 and July 31, 2020 | Australia | Public | 10894 [pre-COVID: 2321, during-COVID: 8573] (I: 6132 [pre-COVID: 5074, during-COVID: 1058]) | Verification of feasibility | Internet CBT | Remote | Medical doctors (and Internet CBT program) | Kessler PHQ GAD | This study assessed the uptake and effectiveness of Internet CBT for symptoms of anxiety and depression during the pandemic and compared outcomes with the 12 months before COVID-19. The Internet CBT course was effective at reducing anxiety, depression, symptom severity, and psychological distress both before and during the pandemic. |
| [25] | Participants were enrolled from | Italy | Patients with COVID-19 | 45 (I: 45) | Verification of feasibility | Tele psychotherapy ("Telecovid Sicilia") | Remote | Psychologist/counselor | SCL-90-R, BDI, ESS, | This study investigated the effects of tele-psychotherapy on emotional well-being |
|  | March to June 2020 |  |  |  |  |  |  |  | HARS. | and psychological distress among patients affected by COVID-19. The intervention improved depression, anxiety, paranoid ideation, and sleep disorders. |
| [26] | Not described in detail; when healthcare professionals fought COVID-19 | China | Healthcare professionals | 4 (I: 4) | Verification of feasibility | Mindfulness-based stress reduction and virtual reality therapy | Virtual reality therapy | Virtual reality | PHQ<br>GAD<br>AIS | This case study examined the effectiveness of virtual reality therapy in the treatment of psychological problems. Four cases of front-line healthcare providers who presented with clinically significant initial psychological problems at the onset of COVID-19 and received virtual reality therapy treatment were described. After sessions, all four cases showed lower scores and improved symptoms for depression, anxiety, psychosomatic symptoms, and sleeping symptoms. |
| [27] | Participants were recruited in April and May 2020; the intervention run from April to November 2020 | Italy | Patients with dementia and their caregivers | 50 (caregiver group: 23, patient group: 25). | RCT | Telephone support | Remote | Psychologist/counselor and master's level licensed social workers | NPI, ZBI, QOL | This study was conducted during the first lockdown period of COVID-19 to provide tele-psychological support to dementia patient caregivers and evaluate its effectiveness by quantifying stress load and quality of life. Caregivers who received telephone support with mood and stress burdens did not experience a worsening of their psychological state during the intervention period. |
| [28] | Not described in detail just mentioned "during | USA | Clinic clients | 4 (I: 4) | Established existing intervention | CBT strategies | Remote | Psychologist/counselor | Fear, sadness, general | This study described the strategy of using CBT for clients with pandemic-related distress at a clinic during the COVID-19 pandemic. A case |
|  | the COVID crisis" |  |  |  |  |  |  |  | distress | of a preliminary intervention was presented in which clinic clients' fear, sadness, and general distress were reduced. |
| [29] | Details were not described (see measures section below; Tennant et al., 2007) | UK | Public | 48 (I: 48) | RCT | ACT | Weekly online semi-interactive, self-help modules and a webinar | Not described | SWEMWBS, CompACT, DASS | This study examined the effectiveness of a guided self-help ACT intervention on well-being for the general population with COVID-related distress and explored the effects of psychological flexibility and distress as well as COVID-19 distress. It showed improvements in well-being, overall psychological flexibility (including the subscales of behavioral awareness and valued action), and general psychological distress (including depression, anxiety, and stress). However, no changes were found for COVID-related distress. |
| [30] | Details were not described | Canada | Temporary migrant live-in caregivers | 46 (I: 18; C: 18) | RCT | ACT | Remote | Not described | DAS, AAQ, CAMS, MSMR | This study evaluated the effectiveness of the online delivery of a six-week psychological intervention based on ACT in reducing psychological distress and promoting resilience among live-in caregivers. The intervention showed improvement in mindful qualities, external resilience, life satisfaction, and accessible support. |
| [31] | Data were collected between January and | USA | Primary caregiver of a child | 12 (I: 12) | Verification of feasibility | Connecting and Reflecting Experience program | Remote | Psychologist/counselor, licensed clinical | PHQ GAD GSRS | The study assessed treatment engagement, acceptability, and psychological distress among participants after the onset of |
|  | August 2020 |  |  |  |  |  |  | psychologists,<br>social workers<br>etc. |  | COVID-19 in the adaptation of telehealth using the Connecting and Reflecting Experience program. Self-reported mood and anxiety symptoms decreased after 20 weeks of telehealth therapy. |

PCL: Post-traumatic Stress Disorder Checklist; HSCL: Hopkins Symptom Checklist; PHQ: Patient Health Questionnaire; GAD: Generalized Anxiety Disorder; HADS: Hospital Anxiety and Depression; IES-R: Impact of Event Scale Revised for Post-traumatic Stress; THERMO: Emotion Thermometer; ISI: Insomnia Severity Index; DAS: Depression, Anxiety, and Stress Scale-21; FCV-19S: Fear of COVID-19 Scale; PWI: Personal Well-being Index; ProQOL: Professional Quality of Life Scale; PSQ: Perceived Sleep Quality; FFMQ: Five Facet Mindfulness Questionnaire; SCS: Self-Compassion Scale; SUDS: Subjective Units of Distress Scale; SCL-90-R: The Symptom Checklist-90-R; BDI: Beck Depression Inventory; ESS: Epworth Sleepiness Scale; HARS: Hamilton Anxiety Rating Scale; AIS: Athens Insomnia Scale; NPI: Neuropsychiatric Inventory; ZBI: burden Interview; QOL: Quality of Life caregiver; SWEMWBS: Warwick-Edinburgh Mental Wellbeing short form; CompACT: the Comprehensive Assessment of Acceptance Commitment Therapy Processes, DASS: Depression Anxiety Stress Scale; AAQ: Acceptance and Action Questionnaire; CAMS: Cognitive and Affective Mindfulness Scale; MSMR: Multi-System Model of Resilience; GSRS: Group Session Rating Scale.

**Figure 2.**
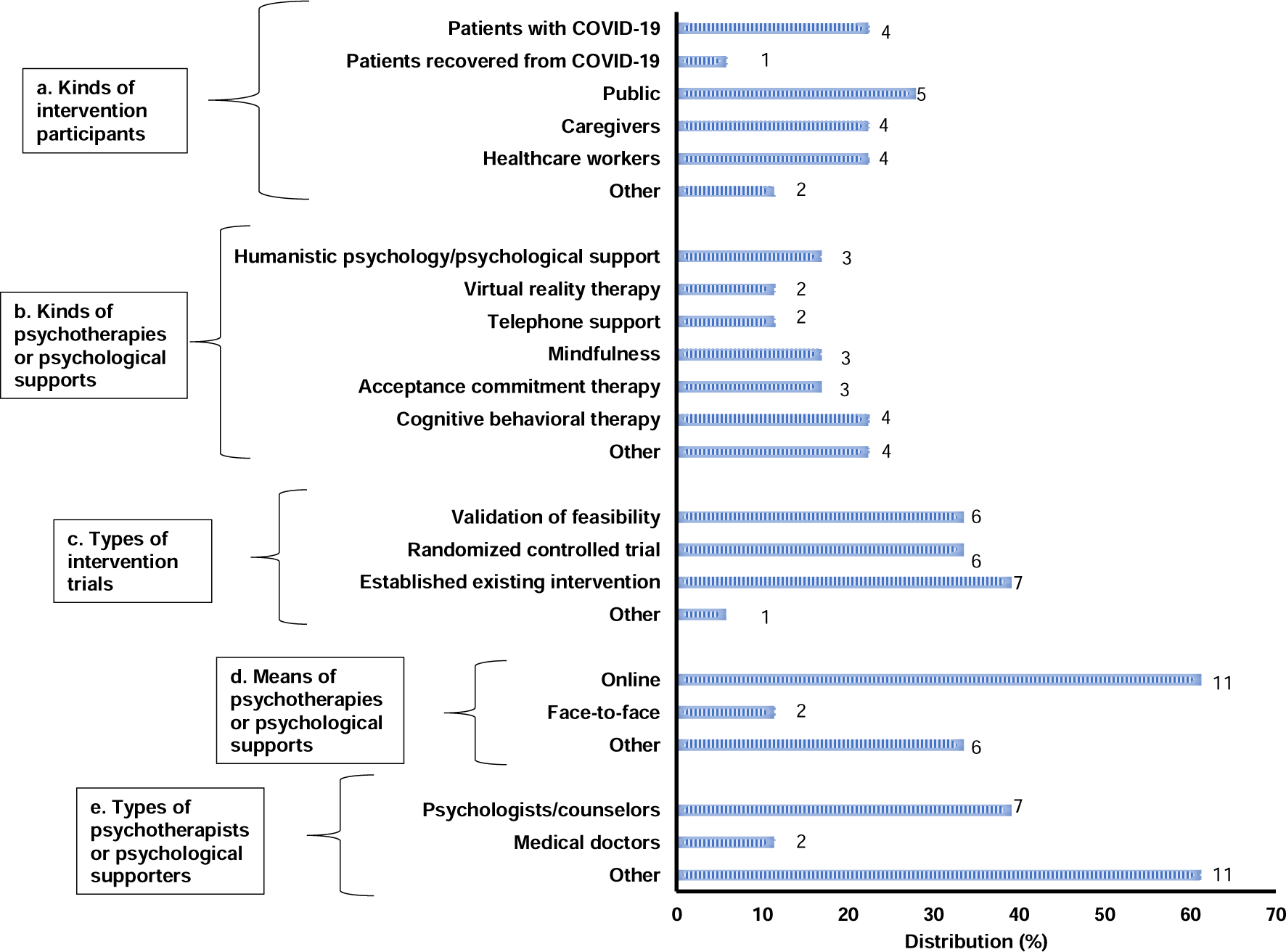
Distribution of the survey items in the subject papers. Bars indicate the distribution (%) of target papers (n = 18). The numbers next to the bars indicate the number of papers that match each item.

a. Types of intervention participants Other (total n = 2; n = 1): clients at the clinic and refugees. The numbers in the figure indicate the number of each item: patients with COVID-19, patients who had recovered from COVID-19, caregivers, healthcare workers, and others.
b. Types of psychotherapies and psychological support Other (total n = 4; n = 1, respectively): Problem Management Plus, EMDR therapy, parenting intervention, and video-based psychotherapy. The numbers in the figure indicate the occurrences of each of item: humanistic psychology/psychological support, virtual reality therapy, telephone support, mindfulness, ACT, CBT, and others.
c. Types of intervention trials Other: exploratory-descriptive design, n = 1
d. Means of psychotherapies and psychological support Other (total n = 6; n = 1): no intervention by any person but by a mobile app, educational video, watching virtual reality therapy, webinars, paper-based system (material), and no description. The numbers in the figure indicate the number of items, online, face-to-face, and others in the articles.
e. Types of psychotherapists and psychological supporters Other (n = 11): trained but non-specialist (n = 1); psycho-oncologist (n = 1); educational program (n = 3); postdoctoral fellows, predoctoral psychology interns, and social workers (n = 1); and not described (n = 5). The numbers in the figure represent the number for each item: psychologists/counselors, medical doctors, and others.

### 3.1 Subjects and participants of the articles included in this study

The subjects and participants in the articles we included in this study were categorized as follows: patients with COVID-19 (n = 4, 22%) [20, 21, 23, 25], patients that had recovered from COVID-19 (n = 1, 6%) [16], citizens (n = 5, 28%) [15, 17, 20, 24, 29], caregivers (n = 4, 22%) [18, 27, 30, 31], and healthcare workers (n = 4, 22%) [19, 20, 22, 26]. Moreover, there were two cases falling under the category of "others": clients at a clinic (n = 1) [28] and refugees (n = 1) [14].

### 3.2 Countries and regions

The distribution of countries and regions was widely distributed across Europe, Central and South Asia, East Asia, North America, and Central America (see Table 2).

**Table 2.** Countries and regions of psychotherapies and psychological support in the reviewed papers.

| **Region** | **Country** | **No. of papers** |
| --- | --- | --- |
| Middle East | Jordan | 1 |
|  | Oman | 1 |
| Europe | Italy | 3 |
|  | United Kingdom | 2 |
| North America | Canada | 2 |
|  | United States | 3 |
| Central America | Mexico | 1 |
| East Asia | China | 3 |
| South Asia | India | 1 |
|  | Singapore | 1 |
|  | Indonesia | 1 |
| Oceania | Australia | 1 |

### 3.3 Types of psychotherapies and psychological support

The distribution of psychotherapies and psychological support was as follows: humanistic psychological support (n = 3, 17%) [16–18], virtual reality therapy (n = 2, 11%) [20, 26], telephone support (n = 2, 11%) [25, 27], mindfulness (n = 3, 17%) [22, 23, 26], ACT (n = 3, 17%) [15, 30, 32], CBT (n = 4, 22%) [15, 24, 28, 32], and others (n = 4, 22%): Problem Management Plus (n = 1) [14], EMDR therapy (n = 1) [19], parenting intervention (n = 1) [31], and video-based psychotherapy (n = 1) [23] (see Figure 2).

### 3.4 Types of intervention trials

The distribution of the types of intervention trials was as follows: validation of feasibility (n = 6, 33%) [20, 23-26, 31], RCTs (n = 6, 33%) [14, 15, 20, 22, 27, 30], established existing intervention (n = 7, 39%) [16, 18-21, 28, 29], and others (n = 1, exploratory-descriptive design, 6%) [17] (see Figure 2).

### 3.5 Means of psychotherapies and psychological support

The distribution of the means of psychotherapies and psychological support was as follows: online (n = 11, 61%) [15, 17-19, 21, 24, 25, 27, 28, 30, 31], face-to-face (n = 2, 11%) [14, 16], and others (n = 6, 33%) [20, 22, 23, 26, 29, 33]. The category of "others" included a paper-based system [21], mobile app [22], educational video [23], watching virtual reality therapy [26], webinar [29], and no description [20] (total n = 6; n = 1, respectively; see Figure 2).

### 3.6 Types of psychotherapists and psychological supporters

The distribution of the types of psychotherapists and psychological supporters was as follows: psychologists/counselors (n = 7, 39%) [15, 16, 19, 25, 27, 28, 31], medical doctors (n = 2, 11%) [21, 24], and others (n =11, 61%). The majority fell under the category of “others” and the breakdown was as follows: trained but non-specialist (n = 1) [14], psycho-oncologist (n = 1) [18], not human but educational program (n = 3) [23, 24, 26], postdoctoral fellows (n = 1) [31], post/predoctoral psychology interns (n = 1) [31], social workers (n =1) [31], and not described (n = 5) [17, 20, 22, 29, 30] (see Figure 2).

### 3.7 Outcomes

The outcomes to measure the effectiveness of the intervention were as follows: depression (Kessler, Beck Depression Inventory), anxiety (Generalized Anxiety Disorder, Hamilton Anxiety Rating Scale), depression and anxiety (Hospital Anxiety and Depression Scale, Hopkins Symptom Checklist, Depression, Anxiety, and Stress Scale), post-traumatic stress (Post-traumatic Stress Disorder Checklist, The Impact of Event Scale – Revised), sleep (Insomnia Severity Index, Athens Insomnia Scale, Epworth Sleepiness Scale, Perceived Sleep Quality), and physical symptoms and general health (Patient Health Questionnaire). The following assessments were performed: Emotion Thermometer, Fear of COVID-19 Scale, Personal Well-being Index, Professional Quality of Life Scale, Subjective Units of Distress Scale, Neuropsychiatric Inventory, Five Facet Mindfulness Questionnaire, Self-Compassion Scale, Digit Span Tests (Forward and Backward), Symptom Checklist-90-R, Zait Burden Interview, Quality of Life Caregiver, Warwick-Edinburgh Mental Wellbeing short form, Comprehensive Assessment of Acceptance Commitment Therapy Processes, Acceptance and Action Questionnaire, Cognitive and Affective Mindfulness Scale, Multi-System Model of Resilience, and Group Session Rating Scale (see Table 1).

## 4. Discussion

In this study, we investigated the types of psychotherapies and psychological support provided during COVID-19, target recipients, and delivery methods employed. This study allowed for the possibility that the methods and means of psychotherapies and psychological support were selected and implemented based on the social situation during the COVID-19 outbreak, considering its unique characteristics as an infectious disease.

As for the target population, a wide range of the general public [15, 17, 20, 24, 29], caregivers [18, 27, 30, 31], healthcare workers [19, 20, 22, 26], COVID-19 patients [20, 21, 23, 25], and patients who had recovered from COVID-19 were found [16]. Although there have been many reports of worsening mental health as a sequela of COVID-19 [34], no research has been conducted on patients with COVID-19 sequelae owing to the difficulty in maintaining a post-discharge connection with patients in the chronic phase of symptoms, such as those in the sequelae (which also leads to difficulties in recruiting study participants). Psychiatric symptoms in the chronic phase are diverse and difficult for both patients and practitioners to identify as sequelae of COVID-19. Alternatively, it is possible that most people with COVID-19 experienced spontaneous remission within approximately one year [35, 36] and it might have been difficult to gather participants for clinical trials. Subsequently, regional diversity was found. This indicates that COVID-19 was a global pandemic that transcended countries and regions. With regard to the types of psychotherapies and psychological support, a new generation of psychotherapy (i.e., ACT and mindfulness) [15, 22, 23, 26, 30, 32] and the use of virtual reality were found [20, 26]. The majority of the means of administration were online interventions rather than traditional face-to-face interventions [15, 17-19, 21, 24, 25, 27, 28, 30, 31]. This can be attributed to the infectious nature of COVID-19. It may also be that sufficient technological innovation had already occurred for non-real face-to-face psychotherapies and psychological support to be provided, and the opportunity for the COVID-19 disaster was taken regarding its implementation. This study’s results indicated that the paradigm of the psychiatric world had also been influenced by the social situation, during the COVID-19 pandemic, but it remains to be seen whether the technologies and support methods rapidly introduced will take hold after the threat of COVID-19 has diminished. Regarding the types of intervention trials, feasibility studies [20, 23-26, 31], RCTs [14, 15, 20, 22, 27, 30], and existing interventions were established [16, 18-21, 28, 29]. Although the majority of the studies in this review provided psychotherapy/support online, only three papers were online RCTs [15, 27, 30]. We were also unable to find any studies that have compared the effectiveness of online and face-to-face psychotherapies and psychological support. Therefore, further research on this topic is required. In this review, interventions by professionals dealing with psychotherapies and psychological support (medical doctors and certified psychotherapists/psychologists) did not account for the majority [15, 16, 19, 21, 24, 25, 27, 28, 31]. This suggests the difficulty in obtaining support from psychiatric professionals, replacement by other professionals or novel technologies, or the current situation in which it is difficult for physicians and psychologists to conduct large-scale intervention trials and publish the results as articles because of their budget and human resources, except in some of the most advanced psychiatric regions. The replacement of psychiatric services with new technology (i.e., apps) offers several advantages to service recipients. Firstly, it enhances accessibility to psychotherapies and psychological support, reducing barriers that may hinder individuals from seeking assistance. Secondly, this approach can lead to the standardization and homogenization of psychotherapies and psychological support, which vary in quality among therapists. Regarding outcomes, a range of mental health-related items were employed, including anxiety, post-traumatic stress, sleep, and the fear of COVID-19 scale, in addition to the primary focus on distress. Notably, most studies utilized multiple outcomes, suggesting a comprehensive assessment of participants’ mental health status to provide robust support.

This study clarified to whom and by what means psychotherapies and psychological support was provided during the COVID-19 pandemic, considering the characteristics of COVID-19 as an infectious disease, the social context of the pandemic, and the methods and means by which it was selected and implemented. However, this review also revealed the following research gaps. Firstly, the implementation of psychotherapies and psychological support for patients with post-COVID-19 sequelae may have been insufficient (patients may have been missed). Secondly, it is necessary to clarify the relative effectiveness of online and face-to-face RCTs during the pandemic. Thirdly, it is unclear whether the technology used during the pandemic will continue to be used. Lastly, research is needed on how physicians, psychologists, and other psychiatric professionals differentiate themselves and coexist with new dehumanizing technologies. These issues need to be closely monitored in the long term.

### 4.1 Limitations

The limitations of this study include the fact that only two search engines were used and only articles written in English were examined. In other words, the data available for the study were limited. Additionally, the selection of psychotherapies and psychological support might have been influenced by factors such as ease of implementation and the availability of evidence-based measures. However, this approach may not fully encompass the complexity and nuances of current clinical situations.

## 5. Conclusion

This study revealed the reality of the countries and regions where psychotherapies and psychological support were implemented during the COVID-19 pandemic, implementers, and means of implementation.

- No studies have been published on the use of psychotherapy for patients suffering from the sequelae of COVID-19. Consequently, patients suffering from post-COVID-19 sequelae might not be receiving adequate support.
- The implementation of psychotherapy was not limited to more advanced regions or countries in psychiatry, indicating the broader reach of these interventions.
- Psychotherapy included a wide range of methods, including new-generation psychotherapy, virtual reality, supportive care, CBT, and online educational content via apps. The rise of new technologies raises questions about the potential replacement of human therapists.
- The majority of psychotherapy interventions were delivered online. However, whether this trend will persist beyond the COVID-19 pandemic remains uncertain. Further research on the differences in effectiveness between online and face-to-face psychotherapy is needed.
- The number of professionals providing psychotherapy, including both physicians and psychologists, appeared to be relatively low.

## Acknowledgement

This work was conducted as part of "The Nippon Foundation - Osaka University Project for Infectious Disease Prevention" and supported by JSPS KAKENHI Grant Number 20K14214 as “Grant-in-Aid for Early-Career Scientists.” We thank Editage [http://www.editage.com] for editing and reviewing the manuscript for English language.

## Ethical Approval

Not required

## Author Approval

All authors have seen and approved the manuscript.

## Competing Interests

No conflicts of interest to declare with respect to this study.

## Funding

This research received funding from two specific grants:

1. JSPS KAKENHI Grant Number 20K14214 as “Grant-in-Aid for Early-Career Scientists.”
2. The Nippon Foundation - Osaka University Project for Infectious Disease Prevention

## Data Availability Statement

All data produced in the present work are contained in the manuscript.

## Statement

During the preparation of this manuscript, the authors used DeepL solely for the purpose of improving English language expression. After using this tool, the authors reviewed and edited the content as needed. Furthermore, the paper was carefully edited by Editage [http://www.editage.com] . The authors take full responsibility for the content of the publication.

## Note

Preliminary results of this study were presented at the 41st Japanese Society for Social Psychiatry Congress and the seminar by SpringX, CiDER, and Knowledge Capital (YouTube: https://youtu.be/TEcwH-IR-38).

## Notes

### Competing Interest Statement

The authors have declared no competing interest.

## References

[1] Taquet M, Geddes JR, Husain M, Luciano S, Harrison PJ. 6-month neurological and psychiatric outcomes in 236 379 survivors of COVID-19: A retrospective cohort study using electronic health records. Lancet Psychiatry. 2021;8:416–27. 10.1016/S2215-0366(21)00084-5.

[2] Engel GL. The need for a new medical model: a challenge for biomedicine. Science. 1977;196:129–36. 10.1126/science.847460.

[3] Huang C, Huang L, Wang Y, Li X, Ren L, Gu X et al. 6-month consequences of COVID-19 in patients discharged from hospital: A cohort study. Lancet. 2021;397:220–32. 10.1016/S0140-6736(20)32656-8.

[4] Nalbandian A, Sehgal K, Gupta A, Madhavan MV, McGroder C, Stevens JS et al. Postacute COVID-19 syndrome. Nat Med. 2021;27:601–15. 10.1038/s41591-021-01283-z.

[5] News, B. Coronavirus: The world in lockdown, in maps and charts. 2020.

[6] Yagihashi M, Murakami M, Kato M, Yamamura A, Miura A, Hirai K. Exploratory study to characterize the individual types of health literacy and beliefs and their associations with infection prevention behaviors amid the COVID-19 pandemic in Japan: a longitudinal study. medRxiv 2023; 2023.2004.2002.23287895.

[7] Murakami M, Hiraishi K, Yamagata M, Nakanishi D, Miura A. Belief in just deserts regarding individuals infected with COVID-19 in Japan and its associations with demographic factors and infection-related and socio-psychological characteristics: a cross-sectional study. PeerJ. 2022;10:e14545. 10.7717/peerj.14545.

[8] Yamagata M, Teraguchi T, Miura A. Effects of pathogen avoidance tendency on infection-prevention behaviors and exclusionary attitudes toward foreigners: A longitudinal study of the COVID-19 outbreak in Japan. Jpn Psychol Res. 2021. 10.1111/jpr.12377.

[9] Xiong J, Lipsitz O, Nasri F, Lui LMW, Gill H, Phan L et al. Impact of COVID-19 pandemic on the mental health of the general population: A systematic review. J Affect Disord. 2020;277:55–64. 10.1016/j.jad.2020.08.001.

[10] Ursano RJW, Lars, Fullerton CS, Raphael B. Textbook of disaster psychiatry, Second edition, textbook of disaster psychiatry. 2nd ed. Cambridge: Cambridge University Press; 2017, p. i–.

[11] Muller AE, Hafstad EV, Himmels JPW, Smedslund G, Flottorp S, Stensland SØ et al. Mental health impact of the Covid-19 pandemic on healthcare workers and interventions to help them: A rapid systematic review. Psychiatry Res. 2020;293:113441. 10.1016/j.psychres.2020.113441.

[12] Bertuzzi V, Semonella M, Bruno D, Manna C, Edbrook-Childs J, Giusti EM et al. Psychological support interventions for healthcare providers and informal caregivers during the COVID-19 pandemic: A systematic literature review Int J Environ Res Public Health. 2021;18:6939. 10.3390/ijerph18136939.

[13] D’Alvano G, Buonanno D, Passaniti C, De Stefano M, Lavorgna L, Tedeschi G et al. Support needs and interventions for family caregivers of patients with amyotrophic lateral sclerosis (ALS): A narrative review with report of telemedicine experiences at the time of COVID-19 pandemic. Brain Sci. 2022;12. 10.3390/brainsci12010049.

[14] Akhtar A, Bawaneh A, Awwad M, Al-Hayek H, Sijbrandij M, Cuijpers P et al. A Longitudinal Study of the Mental Health of Syrian Refugees Before and During the COVID-19 Pandemic Eur J Psychotraumatol. 2021;12:1991651. 10.1080/20008198.2021.1991651.

[15] Al-Alawi M, McCall RK, Sultan A, Al Balushi N, Al-Mahrouqi T, Al Ghailani A et al. Efficacy of a six-week-long therapist-guided online therapy versus self-help internet-based therapy for COVID-19–induced anxiety and depression: Open-label, pragmatic, randomized controlled trial. JMIR Ment Health. 2021;8: e26683. 10.2196/26683.

[16] Bonazza F, Borghi L, di San Marco EC, Piscopo K, Bai F, Monforte AD et al. Psychological outcomes after hospitalization for COVID-19: Data from a multidisciplinary follow-up screening program for recovered patients. Res Psychother. 2020;23:491. 10.4081/ripppo.2020.491.

[17] Briceño Rodríguez MB, Gutiérrez-García RA. Mental health intervention program during the COVID-19 for Mexican adults. Gac Med Caracas. 2022;130 Supl. 3:S540–9. 10.47307/GMC.2022.130.s3.8.

[18] Cheriyalinkal Parambil B, Goswami S, Roy Moulik N, Sonkusare L, Dhamne C, Narula G et al. Psychological distress in primary caregivers of children with cancer during COVID-19 pandemic-A single tertiary care center experience. Psycho-Oncology. 2022;31:253–9. 10.1002/pon.5793.

[19] Fernandez I, Pagani M, Gallina E. Post-traumatic stress disorder among healthcare workers during the COVID-19 pandemic in Italy: Effectiveness of an eye movement desensitization and reprocessing intervention protocol. Front Psychol. 2022;13:964334. 10.3389/fpsyg.2022.964334.

[20] Hatta MH, Sidi H, Siew Koon C, Che Roos NA, Sharip S, Abdul Samad FD et al. Virtual reality (VR) technology for treatment of mental health problems during COVID-19: A systematic review. Int J Environ Res Public Health. 2022;19. 10.3390/ijerph19095389.

[21] He J, Yang L, Pang J, Dai L, Zhu J, Deng Y et al. Efficacy of simplified-cognitive behavioral therapy for insomnia(S-CBTI) among female COVID-19 patients with insomnia symptom in Wuhan mobile cabin hospital. Sleep Breath. 2021;25:2213–9. 10.1007/s11325-021-02350-y.

[22] Keng SL, Chin JWE, Mammadova M, Teo I. Effects of mobile app-based mindfulness practice on healthcare workers: A randomized active controlled trial. Mindfulness. 2022;13:2691–704. 10.1007/s12671-022-01975-8.

[23] Lukman PR, Saputra A, Elvira SD, Heriani, Almasyhur AF, Putri LA et al. Efficacy of video-based psychotherapy in reducing psychological distress of COVID-19 patients treated in isolation ward. Med J Indones. 2021;30:250–255. 10.13181/mji.oa.215473.

[24] Mahoney A, Li I, Haskelberg H, Millard M, Newby JM. The uptake and effectiveness of online cognitive behaviour therapy for symptoms of anxiety and depression during COVID-19. J Affect Disord. 2021;292:197–203. 10.1016/j.jad.2021.05.116.

[25] Maresca G, Formica C, De Cola MC, Lo Buono V, Latella D, Cimino V et al. Care models for mental health in a population of patients affected by COVID-19. J Int Med Res. 2022;50:3000605221097478. 10.1177/03000605221097478.

[26] Pan X, Zhang YC, Ren D, Lu L, Wang YH, Li GX et al. Virtual reality in treatment for psychological problems in first-line health care professionals fighting COVID-19 pandemic: A case series. J Nerv Ment Dis. 2022;210:754–9. 10.1097/NMD.0000000000001531.

[27] Rotondo E, Galimberti D, Mercurio M, Giardinieri G, Forti S, Vimercati R et al. Caregiver tele-assistance for reduction of emotional distress during the COVID-19 pandemic. Psychological support to caregivers of people with dementia: The Italian experience. J Alzheimers Dis. 2022;85:1045–52. 10.3233/JAD-215185.

[28] Sanderson WC, Arunagiri V, Funk AP, Ginsburg KL, Krychiw JK, Limowski AR et al. The nature and treatment of pandemic-related psychological distress. J Contemp Psychother. 2020;50:251–63. 10.1007/s10879-020-09463-7.

[29] Shepherd K, Golijani-Moghaddam N, Dawson DL. AC. ACTing towards better living during COVID-19: The effects of Acceptance and Commitment therapy for individuals affected by COVID-19. J Contextual Behav Sci. 2022;23:98–108. 10.1016/j.jcbs.2021.12.003.

[30] Vahabi M, Pui-Hing Wong JPH, Moosapoor M, Akbarian A, Fung K. Effects of Acceptance and Commitment Therapy (ACT) on mental health and resiliency of migrant live-in caregivers in Canada: Pilot randomized wait list controlled trial. JMIR Form Res. 2022;6: e32136. 10.2196/32136.

[31] Zayde A, Kilbride A, Kucer A, Willis HA, Nikitiades A, Alpert J et al. Connection during COVID-19: Pilot study of a telehealth group parenting intervention. Am J Psychother. 2022;75:67–74. 10.1176/appi.psychotherapy.20210005.

[32] Charney DS. Psychobiological mechanisms of resilience and vulnerability: implications for successful adaptation to extreme stress. Am J Psychiatry. 2004;161:195–216.

[33] Gonzalez Mendez MJ, Ma L, Alvarado R, Ramirez J, Xu KP, Xu HF et al. A multi-center study on the negative psychological impact and associated factors in Chinese healthcare workers 1 year after the COVID-19 initial outbreak. Int J Public Health. 2022;67. 10.3389/ijph.2022.1604979.

[34] Michelen M, Manoharan L, Elkheir N, Cheng V, Dagens A, Hastie C et al. Characterising long COVID: A living systematic review. BMJ Glob Health. 2021;6: e005427. 10.1136/bmjgh-2021-005427.

[35] Zheng YB, Zeng N, Yuan K, Tian SS, Yang YB, Gao N et al. Prevalence and risk factor for long COVID in children and adolescents: A meta-analysis and systematic review. J Infect Public Health. 2023;16:660–72. 10.1016/j.jiph.2023.03.005.

[36] Zeng N, Zhao Y-M, Yan W, Li C, Lu. QD, Liu L, Ni S Y, Mei H, Yuan K, Shi L, Li P, Fan T-T, Yuan J L, Vitiello MV, Kosten T, Kondratiuk AL, Sun H Q, Tang X D, Liu M Y, Lalvani A, Shi J, et al. A systematic review and meta-analysis of the long-term physical and mental sequelae of COVID-19 pandemic: call for research priorities and action. Molecular Psychiatry 2023; 28:423–433

